# Effects of Sports App-Based Interventions on Physical Fitness Outcomes among Adolescents and University Students: A Systematic Review and Meta-Analysis

**DOI:** 10.64898/2026.09.03.26362190

**Authors:** Cong zhou, Wan Ahmad Munsif Bin Wan Pa, Nur Shakila Binti Mazalan, Xiaofen Cao

**Affiliations:** Faculty of Education, Universiti Kebangsaan Malaysia, Bangi, Selangor, Malaysia; Guangxi Vocational & Technical Institute of Industry, Nanning, Guangxi, China; Hunan Agricultural University, Changsha, Hunan, China

## Abstract

**Background:** Sports apps are increasingly used to promote physical activity and support physical education among students. However, their effects on different dimensions of physical fitness and the factors influencing intervention effectiveness remain unclear.

**Objective:** This study aims to evaluate the effects of sports app-based interventions on multiple dimensions of physical fitness among adolescents and students and to examine whether intervention duration, exercise frequency, intensity, and session duration influence intervention effects.

**Methods:** Following PRISMA guidelines, six databases were systematically searched through June 20, 2026. Randomized controlled trials and quasi-experimental studies were included. Risk of bias was assessed, and meta-analyses and subgroup analyses were performed.

**Results:** Twenty moderate-quality studies involving 3,268 adolescents and University students were included. Interventions lasted 6–24 weeks, with 2–7 sessions per week and 20–80 minutes per session. Sports app interventions significantly reduced BMI (MD = −0.41, 95% CI −0.76 to −0.07) and improved cardiorespiratory endurance (SMD = 0.85, 95% CI: 0.35 to 1.35), male pull-ups (MD = 0.74, 95% CI 0.19 to 1.28), female sit-ups (MD = 1.99, 95% CI 1.13 to 2.86), male 1000-m run time (MD = −13.44 s, 95% CI −25.14 to −1.74), female 800-m run time (MD = −9.02 s, 95% CI −13.40 to −4.64), and sit-and-reach performance (MD = 2.01 cm, 95% CI 0.91 to 3.10). Significant subgroup differences by intervention duration were found for cardiorespiratory endurance (P = 0.03) and flexibility (P = 0.01), but no subgroup differences in body mass index were found across the intervention characteristics examined.

**Conclusions:** Sports app interventions can improve multiple dimensions of students’ physical fitness, although current evidence is insufficient to establish an optimal intervention dose. Further high-quality studies need to clarify dose–response relationships and optimize intervention procedures.

## Introduction

Adolescents and University students represent a critical reservoir of future human capital. Their physical fitness not only underpins national development[1]but also shapes their future contributions to society [2]. However, the health status of these populations remains very alarming [3], and approximately 80% of adolescents worldwide fail to meet this recommendation [4]. Adolescent obesity continues to rise [5], accompanied by declines in cardiorespiratory fitness [5] and muscular health [6,7]. The Coronavirus disease 2019 (COVID-19) pandemic further increased sedentary behavior and physical inactivity [8]. Epidemiological studies have shown that higher levels of physical activity and lower levels of sedentary behavior are associated with better physical fitness among adolescents [9]. Consequently, improving the physical fitness of adolescents and University students within educational settings has become a pressing priority in public health and physical education.

Schools provide a critical setting for adolescents and University students to engage in regular exercise through structured curricula, dedicated facilities and qualified teachers [10]. However, traditional physical education is usually standardized, offers limited flexibility and personalization, and shows weak integration with vocational practice [11–14]. With the development of digital technology and the widespread adoption of smart devices, sports apps have emerged as a new approach to promoting physical activity and innovating physical education [15,16]. These apps have become popular because they offer convenience, personalization and interactivity [17]. Features such as goal setting, feedback, activity tracking and personalized recommendations can enhance exercise adherence and motivation [18]. During the COVID-19 pandemic, sports apps also played an important role in supporting active lifestyles and transforming instructional delivery [19–22].

Previous studies have shown that sports app interventions can increase physical activity [23], improve physical fitness and motor skills [24], enhance cardiorespiratory fitness [25] and mental health [26,27], and reduce sport anxiety [28]. However, the findings have been inconsistent. Some experimental studies have reported significantly greater gains in physical activity and physical fitness outcomes in sports app intervention groups than in control groups [29], whereas others have found only limited effects or no significant between-group differences [30,31]. These discrepancies indicate that intervention effects may depend on study design, patterns of app use, and the degree of theoretical grounding [32]. More importantly, despite the growing number of studies on sports apps, systematic evidence on their effects on physical fitness among adolescents and University students remains scarce, and few high-quality evidence syntheses are available.

Therefore, this systematic review and meta-analysis aimed to synthesize the overall effects of sports app–based interventions on five dimensions of physical fitness relevant to human functioning among adolescents and University students: body composition, assessed using BMI; cardiorespiratory endurance; muscular strength; muscular endurance; and flexibility [33]. In addition, the study sought to identify potential moderating effects of intervention and population characteristics and to evaluate the methodological quality of the available evidence, thereby informing future physical education and public health practice.

## Methods

### Protocol Registration

The review was registered with the International Platform of Registered Systematic Review and Meta-analysis Protocols (INPLASY; registration number: INPLASY202620027)[34]. The literature search, review procedures, and reporting followed the Preferred Reporting Items for Systematic Reviews and Meta-Analyses (PRISMA) guidelines[35].

### Search Strategy

Six electronic databases, including PubMed, Web of Science, EBSCOhost, Scopus, Embase, and the Cochrane Library, were searched from inception through June 20, 2026. The search strategies combined controlled vocabulary with free-text terms, as detailed in Table 1. Search terms were grouped into three concepts: population, outcomes, and intervention. Terms within each concept were combined using “OR” and the three concepts were combined using “AND.” All retrieved records were imported into EndNote 21, and duplicates were removed.

**Table 1.**
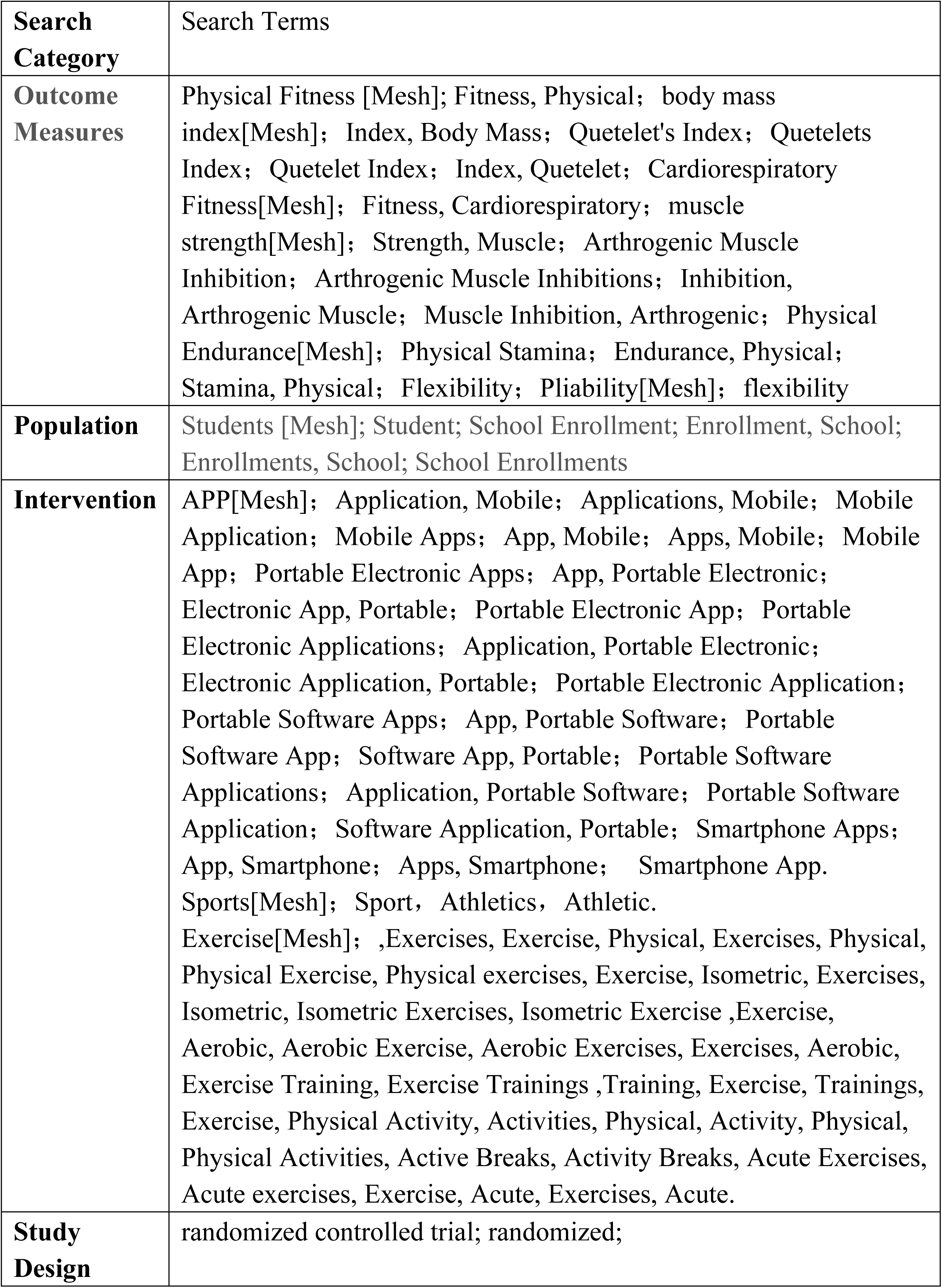
Search Terms.

| **Search Category** | **Search Terms** |
| --- | --- |
| **Outcome Measures** | Physical Fitness [Mesh]; Fitness, Physical; body mass index[Mesh]; Index, Body Mass; Quetelet's Index; Quetelets Index; Quetelet Index; Index, Quetelet; Cardiorespiratory Fitness[Mesh]; Fitness, Cardiorespiratory; muscle strength[Mesh]; Strength, Muscle; Arthrogenic Muscle Inhibition; Arthrogenic Muscle Inhibitions; Inhibition, Arthrogenic Muscle; Muscle Inhibition, Arthrogenic; Physical Endurance[Mesh]; Physical Stamina; Endurance, Physical; Stamina, Physical; Flexibility; Pliability[Mesh]; flexibility |
| **Population** | Students [Mesh]; Student; School Enrollment; Enrollment, School; Enrollments, School; School Enrollments |
| **Intervention** | APP[Mesh]; Application, Mobile; Applications, Mobile; Mobile Application; Mobile Apps; App, Mobile; Apps, Mobile; Mobile App; Portable Electronic Apps; App, Portable Electronic; Electronic App, Portable; Portable Electronic App; Portable Electronic Applications; Application, Portable Electronic; Electronic Application, Portable; Portable Electronic Application; Portable Software Apps; App, Portable Software; Portable Software App; Software App, Portable; Portable Software Applications; Application, Portable Software; Portable Software Application; Software Application, Portable; Smartphone Apps; App, Smartphone; Apps, Smartphone; Smartphone App. Sports[Mesh]; Sport, Athletics, Athletic. Exercise[Mesh]; ,Exercises, Exercise, Physical, Exercises, Physical, Physical Exercise, Physical exercises, Exercise, Isometric, Exercises, Isometric, Isometric Exercises, Isometric Exercise ,Exercise, Aerobic, Aerobic Exercise, Aerobic Exercises, Exercises, Aerobic, Exercise Training, Exercise Trainings ,Training, Exercise, Trainings, Exercise, Physical Activity, Activities, Physical, Activity, Physical, Physical Activities, Active Breaks, Activity Breaks, Acute Exercises, Acute exercises, Exercise, Acute, Exercises, Acute. |
| **Study Design** | randomized controlled trial; randomized; |

### Eligibility Criteria

#### Inclusion Criteria

Studies were included if they met the following criteria: (1) participants were adolescent students aged 10–19 years or University students; (2) the intervention was delivered via a smartphone-based sports or health app; (3) the study employed a randomized controlled trial design; (4) at least one physical fitness outcome was assessed within the five domains of body mass index (BMI), cardiorespiratory fitness, muscular strength, muscular endurance, and flexibility [33]; (5) in light of the heterogeneity of outcome measures across the included studies, physical fitness outcomes were classified into five domains: a. body composition-related outcomes, assessed using BMI; b. cardiorespiratory function, assessed using vital capacity and/or maximal oxygen uptake; c. muscular fitness, assessed using pull-ups in male students and sit-ups in female students; d. endurance running performance, assessed using the 1000-m run in male students and the 800-m run in female students; e. flexibility, assessed using the sit-and-reach test. These field-based measures were considered proxy indicators of the corresponding physical fitness domains rather than direct or interchangeable measures of the underlying physiological constructs; (6) sufficient pre- and post-intervention data, or other data necessary to calculate an effect size, were reported; and (7) the study was an original article published in Chinese or English [33].

#### Exclusion Criteria

Studies were excluded if they met any of the following criteria: (1) duplicate publication; (2) publication in a language other than Chinese or English; (3) classification as a review article, animal study, conference abstract, book chapter, commentary, or dissertation; (4) inclusion of athletes, participants with medical conditions, or participants outside the eligible age range; (5) use of an intervention that was not delivered via a smartphone app or did not target physical activity, exercise promotion, or physical fitness; (6) lack of clearly defined outcome measures or insufficient data for effect-size calculation, with the required data still unavailable after contacting the corresponding author; or (7) use of a study design other than a randomized controlled trial.

#### Data Extraction

Following the literature search, all retrieved records were imported into EndNote 21 and duplicates were removed. The prespecified eligibility criteria were then applied. Two independent reviewers (ZC and WAN) screened the titles and abstracts of all records and subsequently assessed all potentially eligible full-text articles. Disagreements at either stage were resolved in consultation with a third reviewer (SHAKKA) until consensus was reached.

Data from eligible studies were extracted using a prespecified Microsoft Excel form. The extracted information included: (1) author and year of publication; (2) participant characteristics, including age, sex, and sample size; (3) type of sports app; (4) study and intervention characteristics, including intervention duration, exercise frequency, exercise intensity, session duration, randomization procedures, and outcome measures; and (5) intervention results.

#### Risk of Bias Assessment

Risk of bias was evaluated using the Cochrane Risk of Bias tool. The assessment encompassed seven domains: random sequence generation, allocation concealment, blinding of participants and personnel, blinding of outcome assessment, incomplete outcome data, selective reporting, and other potential sources of bias [36]. Each domain was classified as having a low, high, or unclear risk of bias. Discrepancies between reviewers were resolved through discussion with a third independent reviewer (SHAKKA) [36].

#### Statistical Analysis

Outcomes reported by fewer than three studies were summarized narratively. Meta-analysis was conducted when at least three methodologically comparable studies provided extractable pre- and post-intervention data [37]. All statistical analyses were performed using Review Manager 5.4. For continuous outcomes assessed with the same instrument, effect sizes were expressed as mean differences (MDs). When outcomes were measured with different scales or methods, standardized mean differences (SMDs) were computed as Hedges’ g [37]. Means and standard deviations were used to derive effect estimates, which were presented with 95% confidence intervals.

Statistical heterogeneity was evaluated using Cochran’s Q test and the I² statistic. When I² ≤ 50% and P ≥ 0.10, heterogeneity was considered low and a fixed-effect model was adopted. Conversely, when I² > 50% and P < 0.10, heterogeneity was deemed substantial and a random-effects model was applied[37,38].

## Results

### Study Selection

The database search yielded 373 records in total: 19 from PubMed, 49 from Web of Science, 45 from EBSCOhost, 89 from Scopus, 45 from the Cochrane Library, and 126 from Embase. After removal of 113 duplicate records, 260 unique citations remained for screening.

Screening of titles and abstracts excluded 139 records, comprising 6 reviews, 2 articles published in languages other than Chinese or English, 21 non-randomized controlled trials, and 110 records that were irrelevant to the study question. The full texts of the remaining 121 articles were then assessed for eligibility.

Of these, 101 articles were excluded due to ineligible participants (n = 16), ineligible outcomes (n = 56), ineligible interventions (n = 22), or unavailable full texts or required data (n = 7). Consequently, 20 studies met the predefined eligibility criteria and were included in the review. The study selection process is shown in Fig 1.

**Figure 1.**
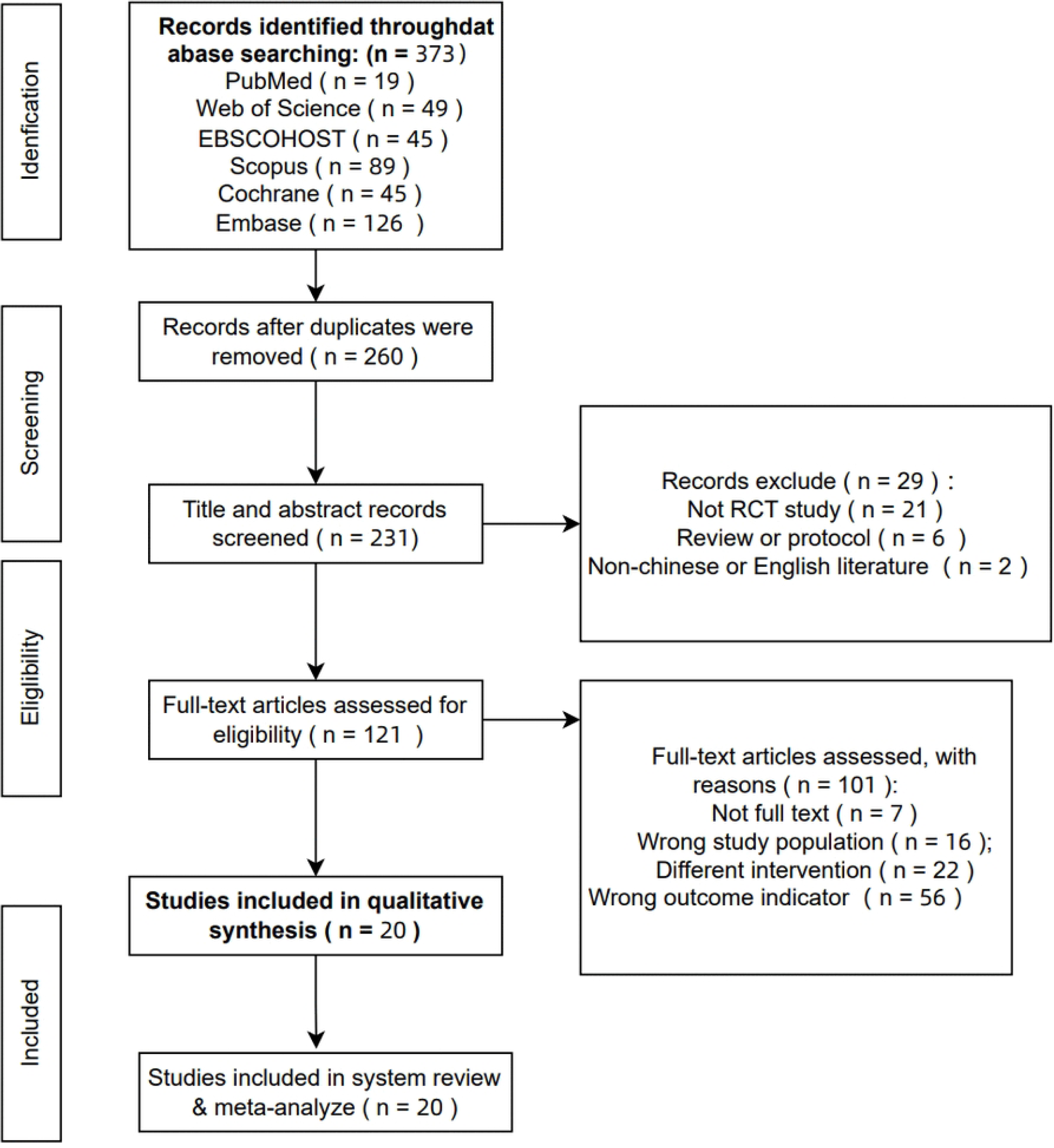
PRISMA flow chart of the study selection process

**Figure 2.**
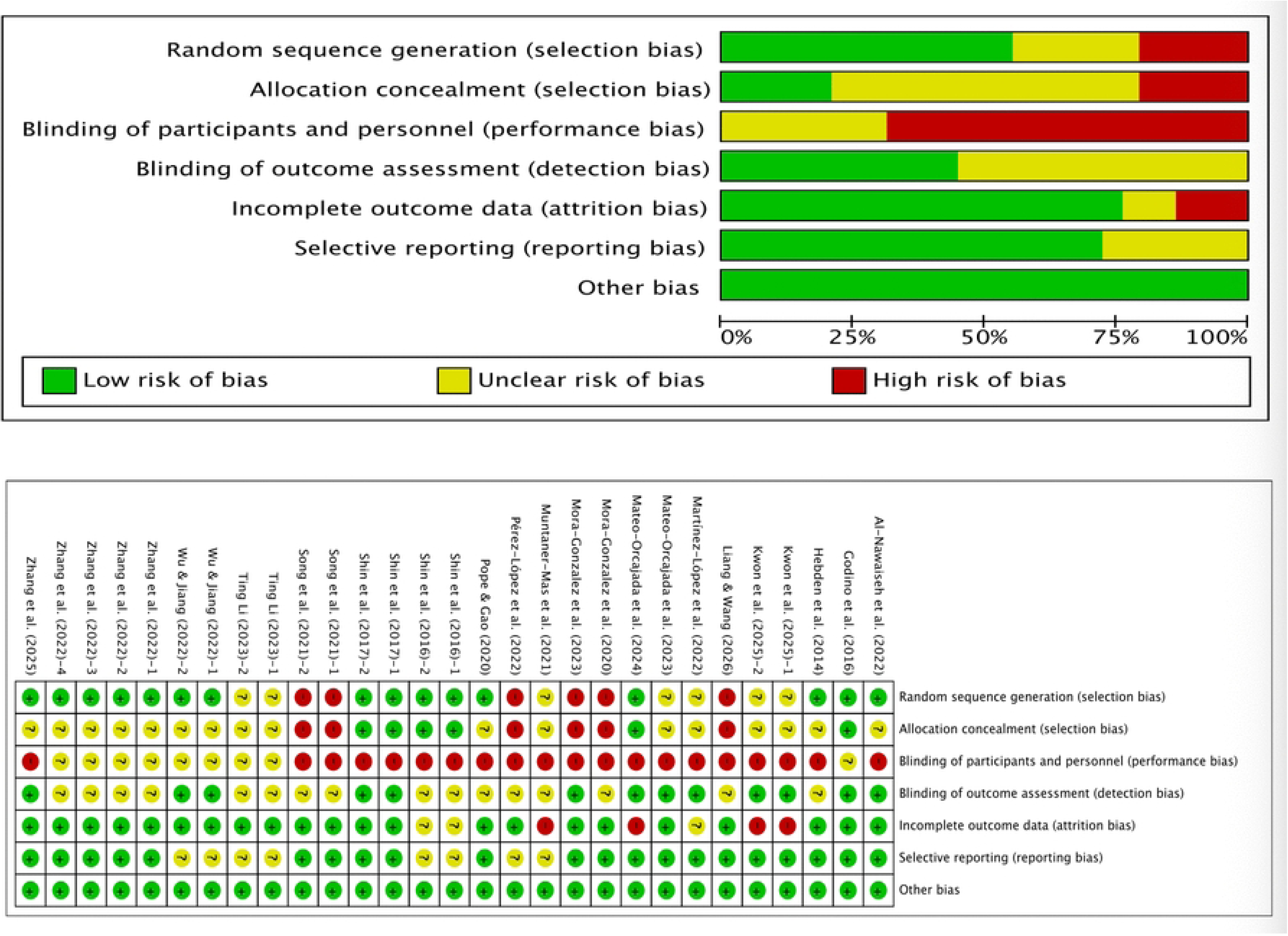
Risk of bias graph and summary

### Characteristics of the Included Studies

#### Characteristics of the Included Studies

Twenty randomized controlled trials published between 2014 and 2026 were included in the review. The samples consisted predominantly of healthy adolescents and University students aged 11–35 years. The evidence base showed marked geographic clustering: seven trials were conducted in Spain[25,39–42], six in China [43–48], four in the United States [49–52], two in South Korea [53,54], and one in Australia [55].

Across all studies, a total of 3,268 participants were enrolled. Individual sample sizes ranged from 33 to 404, yielding an average of approximately 163 participants per trial. Seventeen studies recruited both male and female participants [25,39–42,50–52,55–59], whereas three trials enrolled only male participants [47,53,54]; notably, none focused exclusively on female participants. Four studies disaggregated and reported outcomes by sex [43–46].

In terms of population type, sixteen trials targeted University students [25,40,43–46,50–55,57,58,60], while four focused on adolescents [39,42,49,56].

Intervention duration ranged from 6 to 24 weeks, with the majority spanning 8–16 weeks. Most studies prescribed a minimum of three sessions per week, while several implemented higher frequencies of five to seven sessions per week[49,53,54]. Exercise intensity was generally described as moderate; however, some interventions employed moderate-to-vigorous or high-intensity protocols, and a subset of studies did not report intensity. Individual sessions typically lasted 20–60 minutes. Certain interventions mandated sessions of at least 40 minutes [42,55] whereas a few studies adopted shorter 10–20-minute sessions [40,46]. Detailed study characteristics are presented in Table 2.

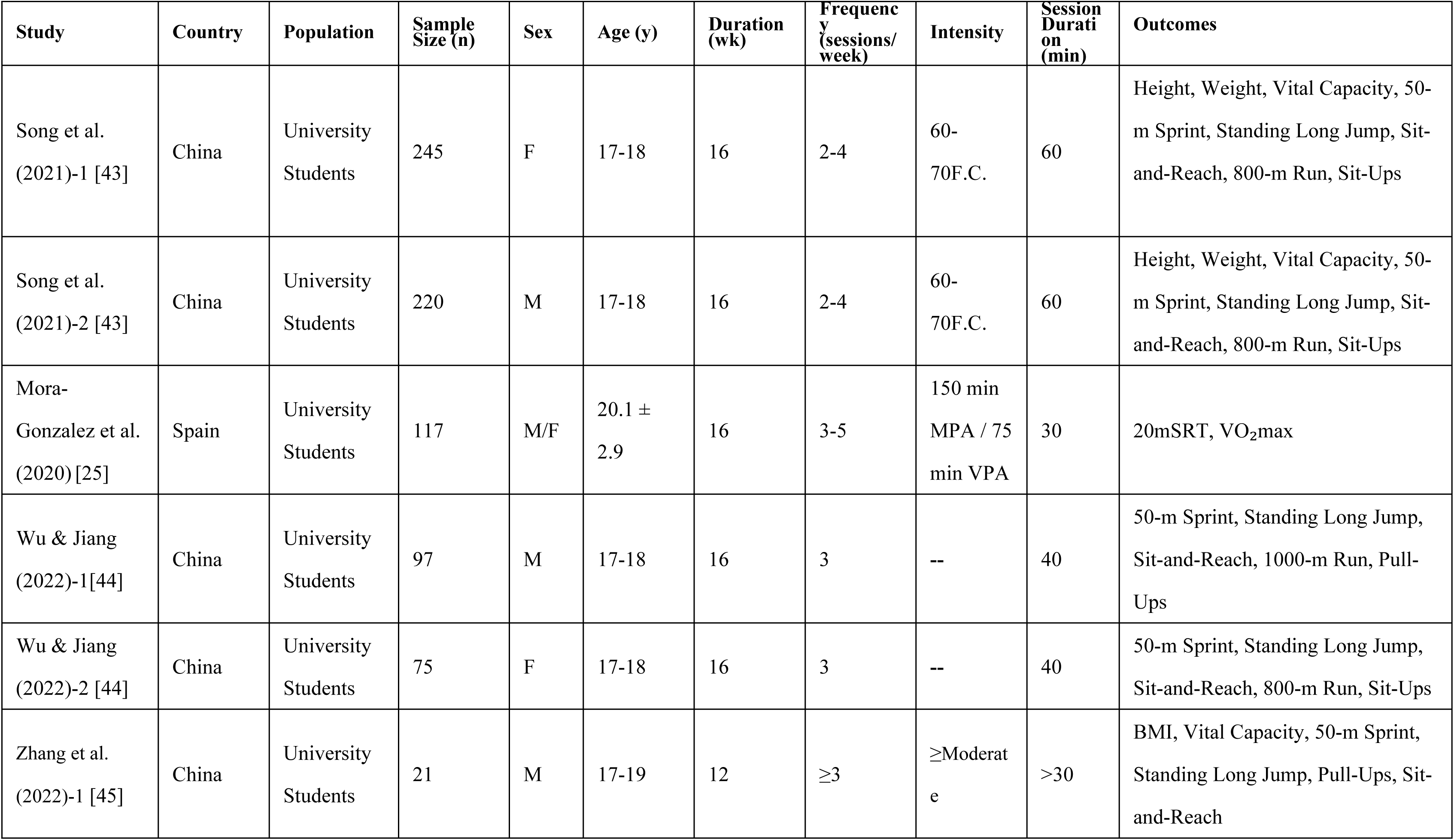
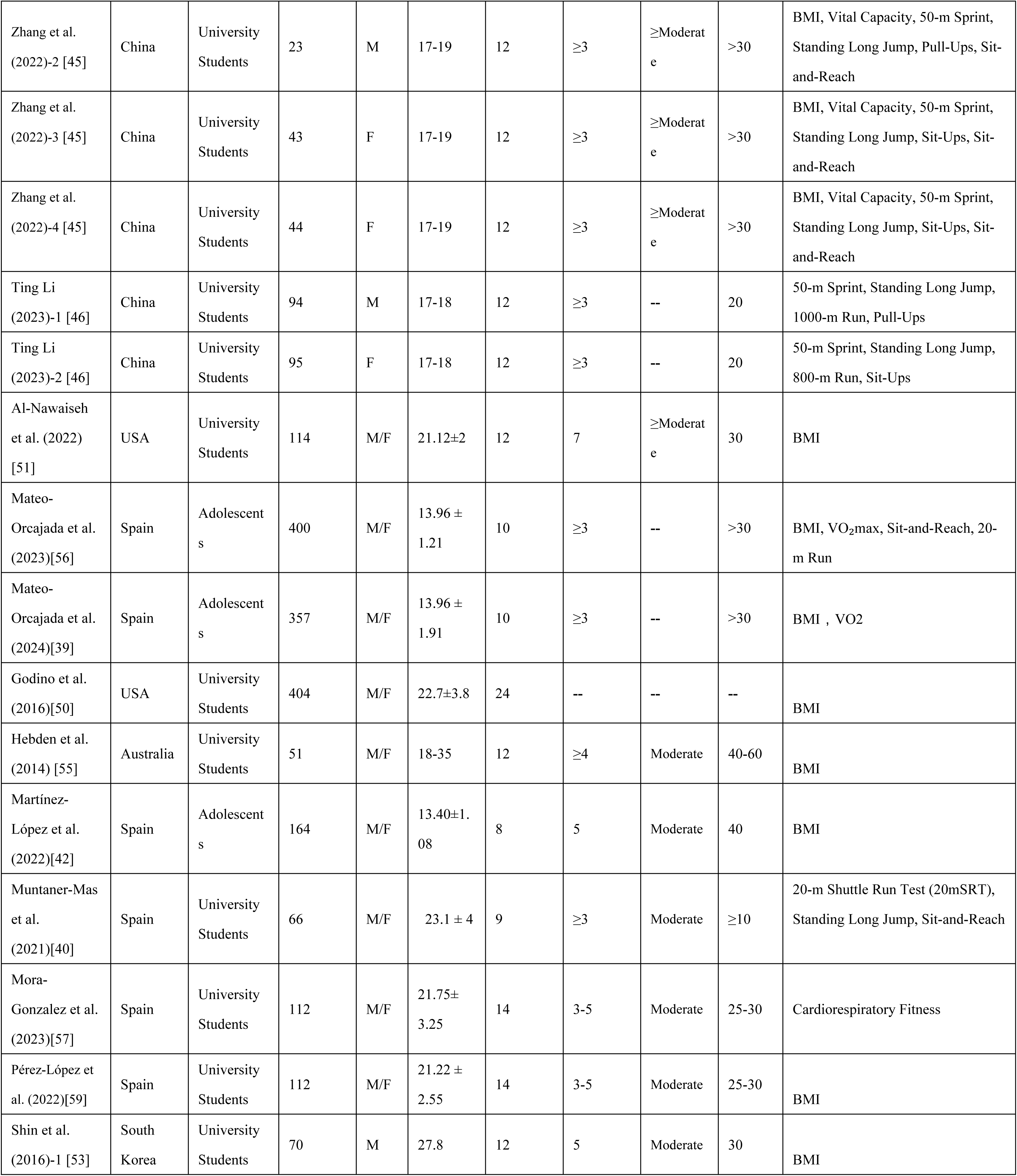
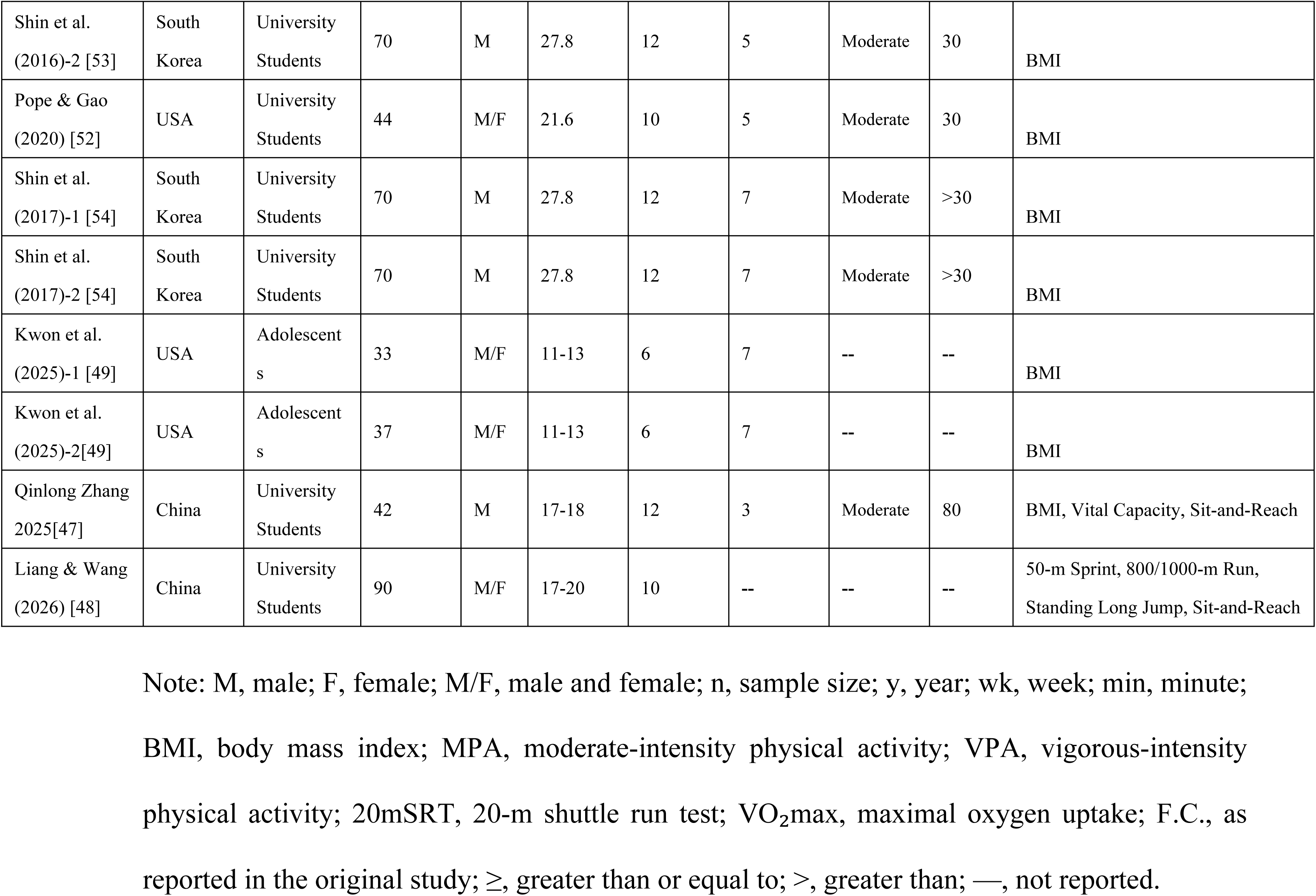

| Study | Country | Population | Sample Size (n) | Sex | Age (y) | Duration (wk) | Frequency (sessions/week) | Intensity | Session Duration (min) | Outcomes |
| --- | --- | --- | --- | --- | --- | --- | --- | --- | --- | --- |
| Song et al. (2021)-1 [43] | China | University Students | 245 | F | 17-18 | 16 | 2-4 | 60-70F.C. | 60 | Height, Weight, Vital Capacity, 50-m Sprint, Standing Long Jump, Sit-and-Reach, 800-m Run, Sit-Ups |
| Song et al. (2021)-2 [43] | China | University Students | 220 | M | 17-18 | 16 | 2-4 | 60-70F.C. | 60 | Height, Weight, Vital Capacity, 50-m Sprint, Standing Long Jump, Sit-and-Reach, 800-m Run, Sit-Ups |
| Mora-Gonzalez et al. (2020)[25] | Spain | University Students | 117 | M/F | 20.1 ± 2.9 | 16 | 3-5 | 150 min MPA / 75 min VPA | 30 | 20mSRT, VO<sub>2</sub>max |
| Wu & Jiang (2022)-1[44] | China | University Students | 97 | M | 17-18 | 16 | 3 | -- | 40 | 50-m Sprint, Standing Long Jump, Sit-and-Reach, 1000-m Run, Pull-Ups |
| Wu & Jiang (2022)-2 [44] | China | University Students | 75 | F | 17-18 | 16 | 3 | -- | 40 | 50-m Sprint, Standing Long Jump, Sit-and-Reach, 800-m Run, Sit-Ups |
| Zhang et al. (2022)-1 [45] | China | University Students | 21 | M | 17-19 | 12 | ≥3 | ≥Moderate | >30 | BMI, Vital Capacity, 50-m Sprint, Standing Long Jump, Pull-Ups, Sit-and-Reach |
| Zhang et al.<br>(2022)-2 [45] | China | University Students | 23 | M | 17-19 | 12 | $\geq 3$ | ≥Moderate | >30 | BMI, Vital Capacity, 50-m Sprint, Standing Long Jump, Pull-Ups, Sit-and-Reach |
| Zhang et al.<br>(2022)-3 [45] | China | University Students | 43 | F | 17-19 | 12 | $\geq 3$ | ≥Moderate | >30 | BMI, Vital Capacity, 50-m Sprint, Standing Long Jump, Sit-Ups, Sit-and-Reach |
| Zhang et al.<br>(2022)-4 [45] | China | University Students | 44 | F | 17-19 | 12 | $\geq 3$ | ≥Moderate | >30 | BMI, Vital Capacity, 50-m Sprint, Standing Long Jump, Sit-Ups, Sit-and-Reach |
| Ting Li<br>(2023)-1 [46] | China | University Students | 94 | M | 17-18 | 12 | $\geq 3$ | -- | 20 | 50-m Sprint, Standing Long Jump, 1000-m Run, Pull-Ups |
| Ting Li<br>(2023)-2 [46] | China | University Students | 95 | F | 17-18 | 12 | $\geq 3$ | -- | 20 | 50-m Sprint, Standing Long Jump, 800-m Run, Sit-Ups |
| Al-Nawaiseh et al. (2022) [51] | USA | University Students | 114 | M/F | 21.12±2 | 12 | 7 | ≥Moderate | 30 | BMI |
| Mateo-Orcajada et al. (2023)[56] | Spain | Adolescents | 400 | M/F | 13.96 ± 1.21 | 10 | $\geq 3$ | -- | >30 | BMI, VO<sub>2</sub>max, Sit-and-Reach, 20-m Run |
| Mateo-Orcajada et al. (2024)[39] | Spain | Adolescents | 357 | M/F | 13.96 ± 1.91 | 10 | $\geq 3$ | -- | >30 | BMI , VO2 |
| Godino et al. (2016)[50] | USA | University Students | 404 | M/F | 22.7±3.8 | 24 | -- | -- | -- | BMI |
| Hebden et al. (2014) [55] | Australia | University Students | 51 | M/F | 18-35 | 12 | $\geq 4$ | Moderate | 40-60 | BMI |
| Martínez-López et al. (2022)[42] | Spain | Adolescents | 164 | M/F | 13.40±1.08 | 8 | 5 | Moderate | 40 | BMI |
| Muntaner-Mas et al. (2021)[40] | Spain | University Students | 66 | M/F | 23.1 ± 4 | 9 | $\geq 3$ | Moderate | $\geq 10$ | 20-m Shuttle Run Test (20mSRT), Standing Long Jump, Sit-and-Reach |
| Mora-Gonzalez et al. (2023)[57] | Spain | University Students | 112 | M/F | 21.75± 3.25 | 14 | 3-5 | Moderate | 25-30 | Cardiorespiratory Fitness |
| Pérez-López et al. (2022)[59] | Spain | University Students | 112 | M/F | 21.22 ± 2.55 | 14 | 3-5 | Moderate | 25-30 | BMI |
| Shin et al. (2016)-1 [53] | South Korea | University Students | 70 | M | 27.8 | 12 | 5 | Moderate | 30 | BMI |
| Shin et al.<br>(2016)-2 [53] | South Korea | University Students | 70 | M | 27.8 | 12 | 5 | Moderate | 30 | BMI |
| Pope & Gao<br>(2020) [52] | USA | University Students | 44 | M/F | 21.6 | 10 | 5 | Moderate | 30 | BMI |
| Shin et al.<br>(2017)-1 [54] | South Korea | University Students | 70 | M | 27.8 | 12 | 7 | Moderate | >30 | BMI |
| Shin et al.<br>(2017)-2 [54] | South Korea | University Students | 70 | M | 27.8 | 12 | 7 | Moderate | >30 | BMI |
| Kwon et al.<br>(2025)-1 [49] | USA | Adolescents | 33 | M/F | 11-13 | 6 | 7 | -- | -- | BMI |
| Kwon et al.<br>(2025)-2[49] | USA | Adolescents | 37 | M/F | 11-13 | 6 | 7 | -- | -- | BMI |
| Qinlong Zhang<br>2025[47] | China | University Students | 42 | M | 17-18 | 12 | 3 | Moderate | 80 | BMI, Vital Capacity, Sit-and-Reach |
| Liang & Wang<br>(2026) [48] | China | University Students | 90 | M/F | 17-20 | 10 | -- | -- | -- | 50-m Sprint, 800/1000-m Run, Standing Long Jump, Sit-and-Reach |

Note: M, male; F, female; M/F, male and female; n, sample size; y, year; wk, week; min, minute; BMI, body mass index; MPA, moderate-intensity physical activity; VPA, vigorous-intensity physical activity; 20mSRT, 20-m shuttle run test; VO<sub>2</sub>max, maximal oxygen uptake; F.C., as reported in the original study; ≥, greater than or equal to; >, greater than; —, not reported.

### Risk of Bias

The risk-of-bias assessment showed that 10studies (50.0%) had a low risk of bias for random sequence generation [39,44,45,47,50–55]. while only 4 studies (20.0%) had a low risk for allocation concealment [39,50,53,54].

Blinding of participants and personnel was the primary source of bias, with 16 studies (80.0%) rated as having a high risk [25,39,40,42,43,47–49,51–57,59]. For blinding of outcome assessors, 10 studies (50.0%) were rated as having a low risk [39,42,44,47,49–51,54,56,57].

Regarding incomplete outcome data, 15 studies (75.0%) had a low risk of bias [25,43–48,50–52,54–57,59], 3 (15.0%) had a high risk [39,40,49], and 2 (10.0%) had an unclear risk [42,53]. For selective reporting, 15 studies (75.0%) were rated as having a low risk[25,39,42,43,45,47–52,54–57]. All 20 studies had a low risk of other bias. Overall, the main methodological concerns were related to blinding of participants and personnel and allocation concealment.

### Sensitivity Analysis

A leave-one-out sensitivity analysis was performed using Review Manager 5.4 to assess the robustness of the findings. Sequential exclusion of individual studies did not materially alter the pooled effect estimates or their 95% confidence intervals, and the direction and statistical significance of the effects remained unchanged. These findings indicated that the meta-analysis results were robust.

### Assessment of Publication Bias

Publication bias for each physical fitness outcome was evaluated using funnel plots (Fig 3). Across outcomes, the study-specific estimates appeared relatively evenly dispersed, and no pronounced systematic asymmetry emerged. For BMI and sit-and-reach, the data points clustered around the pooled effect estimate and were approximately balanced on both sides of the funnel, indicating a low likelihood of publication bias.

**Figure 3.**
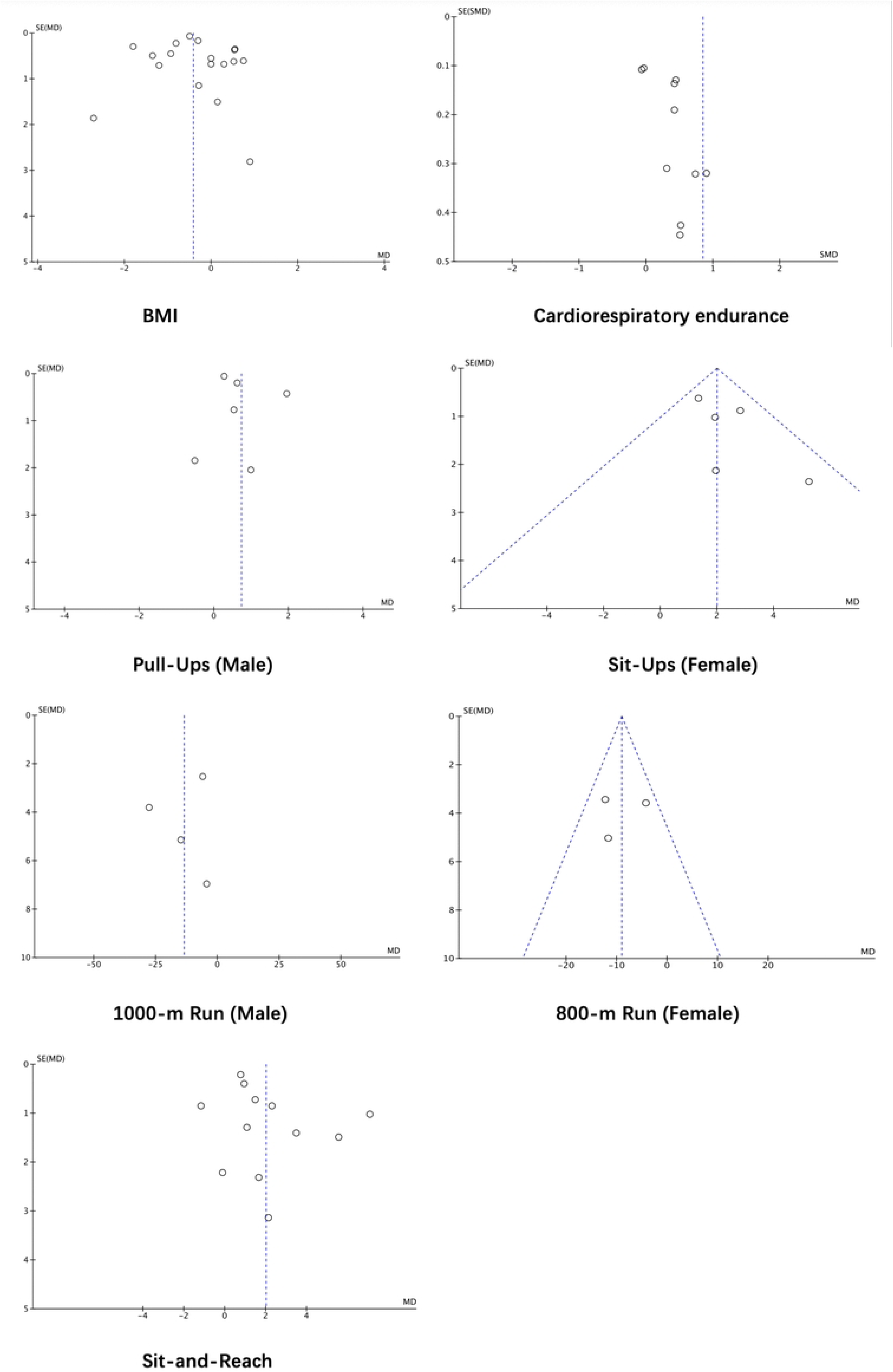
Funnel plot of physical fitness

**Figure 4.**
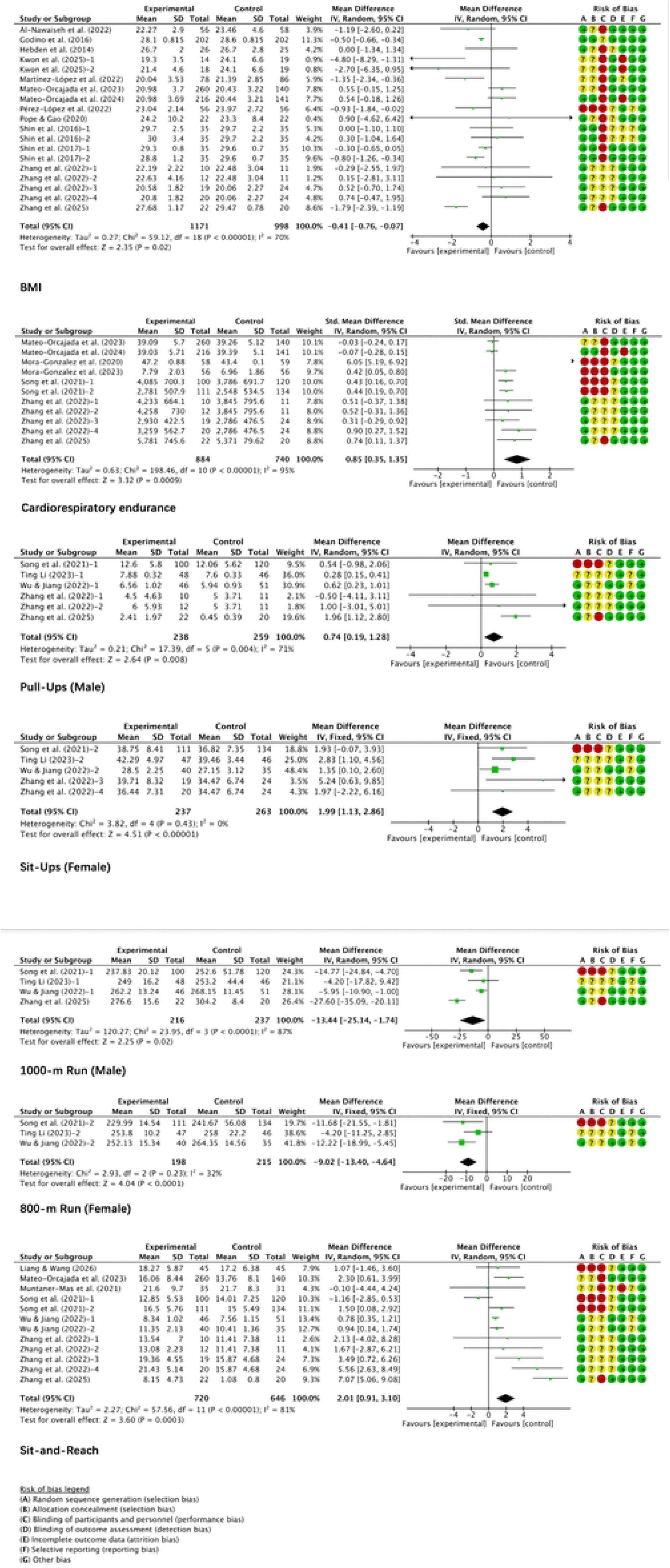
Forest plots of physical fitness

Nonetheless, gaps were evident in the lower portion of several plots, potentially reflecting a relative scarcity of small-sample studies, particularly those reporting smaller or non-significant effects. The funnel plot for cardiorespiratory endurance exhibited noticeable asymmetry, which may have been driven by variability in outcome definitions, testing protocols, and between-study heterogeneity.

For the 1000-m run in males, 800-m run in females, pull-ups in males, and sit-ups in females, the number of eligible studies was limited, and the data points were sparse, precluding a reliable visual assessment of funnel plot symmetry and publication bias. Taken together, the analyses did not reveal any clear or systematic publication bias across the physical fitness outcomes, suggesting that the pooled effect estimates were relatively robust.

### Meta-Analysis and Pooled Effect Estimates

The meta-analysis indicated that sports app interventions significantly enhanced multiple physical fitness indicators in adolescents and University Students. For body composition, the random-effects model showed that BMI was significantly lower in the intervention group than in the control group (MD = −0.41, 95% CI [−0.76, −0.07], P = 0.02; I² = 70%).

In terms of cardiorespiratory function, sports app interventions yielded a significant improvement relative to control conditions (SMD = 0.85, 95% CI [0.35, 1.35], P = 0.0009; I² = 95%). The substantial heterogeneity across studies may have been attributable to variation in cardiorespiratory endurance test protocols, differences in measurement units, and inconsistencies in the handling of effect sizes.

Muscular fitness outcomes improved following sports app interventions. Among male students, pull-up performance increased significantly (MD = 0.74, 95% CI [0.19, 1.28], P = 0.008; I² = 71%), although this effect was accompanied by substantial between-study heterogeneity. Among female students, sit-up performance also increased significantly (MD = 1.99, 95% CI [1.13, 2.86], P < 0.00001; I² = 0%), and no heterogeneity was detected.

Endurance running performance likewise improved. Completion times decreased significantly for the male 1000-m run (MD = −13.44 s, 95% CI [−25.14, −1.74], P = 0.02; I² = 87%) and for the female 800-m run (MD = −9.02 s, 95% CI [−13.40, −4.64], P < 0.0001; I² = 32%). With respect to flexibility, sports app interventions were associated with a significant improvement in sit-and-reach performance (MD = 2.01 cm, 95% CI [0.91, 3.10], P = 0.0003; I² = 81%). Taken together, these pooled estimates indicated that sports app interventions favored improvements in BMI, cardiorespiratory function, muscular fitness, endurance running performance, and flexibility. Nevertheless, the presence of substantial heterogeneity for several outcomes suggested considerable variation in the magnitude of effects across studies. In addition, the small number of studies contributing to some pooled analyses likely limited the precision of the summary estimates.

### Subgroup analysis

To further evaluate whether the effectiveness of sports app–based interventions differed by exercise prescription parameters, we performed subgroup analyses for BMI, cardiorespiratory function, and sit-and-reach performance stratified by intervention duration, exercise frequency, exercise intensity, and session duration. Subgroup analyses were not performed for the male 1000-m run, female 800-m run, male pull-ups, or female sit-ups because these outcomes were reported in an insufficient number of studies. The detailed results of these subgroup analyses are presented in Table 3.

**Table 3.**
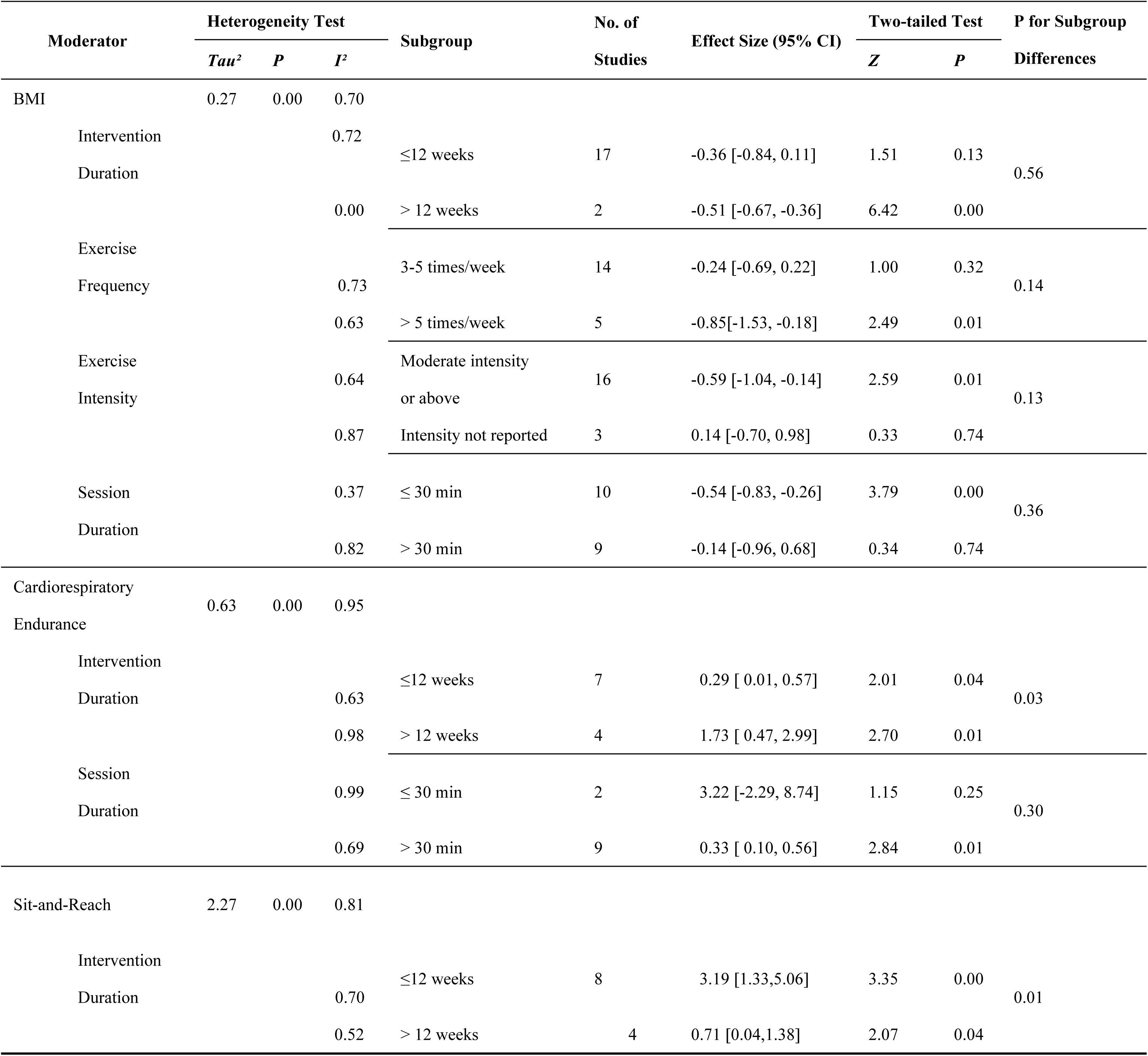
Subgroup Analyses of the Effects of Sports App Interventions on Physical Fitness Outcomes.

#### Subgroup Analysis of BMI

Subgroup analysis by intervention duration indicated that BMI reduction was not statistically significant for interventions lasting ≤12 weeks (MD = −0.36, 95% CI [−0.84, 0.11], P = 0.13; I² = 72%), whereas interventions extending beyond 12 weeks significantly reduced BMI (MD = −0.51, 95% CI [−0.67, −0.36], P < 0.00001; I² = 0%). Nonetheless, the between-subgroup comparison was not statistically significant (P = 0.56), indicating that intervention duration did not moderate the effect on BMI.

Similarly, subgroup analysis by exercise frequency showed that interventions delivered three to five times per week did not significantly change BMI (MD = −0.24, 95% CI [−0.69, 0.22], P = 0.32; I² = 73%), whereas interventions administered more than five times per week significantly reduced BMI (MD = −0.85, 95% CI [−1.53, −0.18], P = 0.01; I² = 63%). However, the difference between these frequency subgroups was not statistically significant (P = 0.14), so the available evidence did not support a greater effect of higher exercise frequency.

Interventions delivered at moderate or higher intensity significantly reduced BMI (MD = −0.59, 95% CI [−1.04, −0.14], P = 0.01; I² = 64%), whereas interventions for which exercise intensity was not reported produced no significant change in BMI (MD = 0.14, 95% CI [−0.70, 0.98], P = 0.74; I² = 87%). However, the between-subgroup comparison was not statistically significant (P = 0.13). Consequently, although moderate-to-vigorous interventions appeared more effective, these findings did not support exercise intensity as a definitive moderating factor.

Subgroup analysis by session duration demonstrated a significant reduction in BMI for sessions lasting ≤30 minutes (MD = −0.54, 95% CI [−0.83, −0.26], P = 0.0001; I² = 37%), whereas sessions longer than 30 minutes yielded no significant effect (MD = −0.14, 95% CI [−0.96, 0.68], P = 0.74; I² = 82%). The between-subgroup comparison was likewise not statistically significant (P = 0.36), so the available data did not provide sufficient evidence that session duration moderated the effect on BMI.

Taken together, certain subgroups defined by intervention duration, exercise frequency, exercise intensity, and session duration exhibited significant reductions in BMI. Nonetheless, none of the four tests for subgroup differences reached statistical significance, indicating that there was insufficient evidence that these intervention parameters modified the impact of sports apps on BMI.

#### Subgroup Analysis of Cardiorespiratory Function

Subgroup analysis by intervention duration indicated that programs lasting ≤12 weeks significantly improved cardiorespiratory function (SMD = 0.29, 95% CI [0.01, 0.57], P = 0.04; I² = 63%). Interventions extending beyond 12 weeks also yielded significant gains (SMD = 1.73, 95% CI [0.47, 2.99], P = 0.007; I² = 98%). The difference between these subgroups was statistically significant (P = 0.03), suggesting that the magnitude of improvement in cardiorespiratory function varied with intervention duration. Nevertheless, because between-study heterogeneity in the >12-week subgroup was substantial, this result required confirmation in future investigations.

Subgroup analysis by session duration indicated that the pooled effect was not statistically significant for sessions lasting ≤30 minutes (SMD = 3.22, 95% CI [−2.29, 8.74], P = 0.25; I² = 99%). In contrast, sessions lasting >30 minutes significantly improved cardiorespiratory function (SMD = 0.33, 95% CI [0.10, 0.56], P = 0.005; I² = 69%). Nevertheless, the between-subgroup difference was not statistically significant (P = 0.30), so the data provided insufficient evidence that session duration moderated the effect of sports apps on cardiorespiratory function.

Subgroup analyses based on exercise frequency and intensity were not conducted because the intervention protocols were highly clustered within these ranges. Most trials prescribed exercise at least three times per week at moderate intensity, leaving too little between-study variability to support informative subgroup comparisons. Overall, longer intervention duration appeared to be associated with greater improvements in cardiorespiratory function. Interventions that yielded beneficial effects typically combined moderate-intensity exercise with sessions of at least 30 minutes and a minimum frequency of three sessions per week. However, apart from intervention duration, the available evidence did not indicate independent moderating effects of the other exercise prescription parameters. Additional high-quality studies are needed to substantiate these observations.

#### Subgroup Analysis of Sit-and-Reach Performance

Subgroup analysis by intervention duration indicated that interventions lasting ≤12 weeks were associated with a significant improvement in sit-and-reach performance (MD = 3.19, 95% CI [1.33, 5.06], P = 0.0008; I² = 70%). Interventions extending beyond 12 weeks likewise yielded a statistically significant benefit (MD = 0.71, 95% CI [0.04, 1.38], P = 0.04; I² = 52%). The between-subgroup difference reached statistical significance (P = 0.01), implying that intervention duration may have moderated the impact of sports apps on flexibility. Notably, the estimated effect was larger for interventions of ≤12 weeks than for those exceeding 12 weeks.

Because most intervention protocols clustered within similar parameter’s moderate-intensity exercise, sessions lasting ≥30 minutes, and a frequency of ≥3 sessions per week) no significant differences were detected with respect to exercise frequency, intensity, or session duration. Nevertheless, the overall findings showed that sports app interventions significantly improved flexibility.

Overall, intervention duration may have moderated the effects of sports apps on cardiorespiratory function and sit-and-reach performance. In contrast, the effects on BMI were not significantly moderated by intervention duration, exercise frequency, exercise intensity, or session duration. Given the small number of studies and substantial heterogeneity in some subgroups, these findings require confirmation through further high-quality research.

## Discussion

This systematic review and meta-analysis showed that sports app interventions generally produced beneficial effects on students’ physical fitness, with particularly notable improvements in body composition-related indicators, cardiorespiratory function, muscular fitness, endurance running performance, and flexibility. The magnitude of these effects, however, and the consistency of findings across studies differed substantially among outcome domains. Considerable heterogeneity emerged for BMI, overall cardiorespiratory function, and sit-and-reach performance, whereas heterogeneity was comparatively lower for several muscular fitness indices and endurance running outcomes. Taken together, these results suggested that the impact of sports app interventions was likely shaped by a constellation of factors, including the underlying exercise prescriptions, specific app functionalities, participant characteristics, and the methods used to assess outcomes.

### Effects of Sports Apps on BMI Among Adolescents and University Students

The meta-analysis indicated that sports app–based interventions significantly reduced BMI (MD = −0.41, 95% CI [−0.76, −0.07], P = 0.02), although substantial heterogeneity existed across studies (I² = 70%). At the level of individual trials, the results of Godino et al. [50], Martínez-López et al.[42], Pérez-López et al.[59], Shin et al.[54], and Zhang et al.[47] generally favored the intervention arms. Among these, Zhang et al. [47] documented a particularly marked reduction in BMI among overweight and obese University Students. By contrast, Mateo-Orcajada et al. [39,56]and Shin et al.[53]reported relatively limited effects, indicating that the effects of sports apps on BMI were not consistent across interventions. The mHealth randomized controlled trial conducted by Zhang et al.[47] focused specifically on overweight and obese University Students and evaluated body composition and physical fitness as primary outcomes. The larger potential for improvement in this high-risk group suggested that baseline weight status may have shaped the magnitude of the observed intervention effect.

Although several subgroups exhibited notable reductions in BMI, specifically interventions lasting >12 weeks, those delivered more than five times per week, those of at least moderate intensity, and those with session durations ≤30 minutes, none of the formal tests for between-subgroup differences reached statistical significance. The P values were 0.56 for intervention duration, 0.14 for exercise frequency, 0.13 for exercise intensity, and 0.36 for session length. On this basis, the evidence did not demonstrate that any single exercise-prescription parameter independently moderated the effect on BMI. Rather, the observed changes in BMI likely reflected the integrated influence of total exercise volume, baseline body composition, intervention adherence, and other concomitant lifestyle factors.

### Effects of Sports Apps on Cardiorespiratory Function Among Adolescents and University Students

Sports app interventions produced a significant overall effect on cardiorespiratory function (SMD = 0.85, 95% CI [0.35, 1.35], P = 0.0009), although the degree of between-study heterogeneity was substantial (I² = 95%). Within the body of included trials, Mora-Gonzalez et al. [25] reported a comparatively large beneficial effect. Consistent with this pattern, Song et al.[43], Mora-Gonzalez et al.[57], and Zhang et al. [47] likewise reported results that generally favored the intervention groups, whereas Mateo-Orcajada et al. [39,56] documented notably smaller effect sizes.

The considerable heterogeneity may have been partly attributable to variation in the instruments used to evaluate cardiorespiratory function. Across the included studies, investigators employed vital capacity along with a range of other aerobic endurance-related tests, each capturing distinct dimensions of cardiorespiratory physiology. Intervention duration and exercise dose likewise differed among studies. For instance, Mora-Gonzalez et al. [57]implemented a 14-week intervention, Shin et al. [53,54] conducted 12-week interventions, and several other studies adopted protocols lasting 10–12 weeks. Collectively, these discrepancies in assessment tools and intervention length underscored substantial differences in training exposure among the studies.

Subgroup analyses indicated that interventions of both ≤12 weeks and >12 weeks significantly improved cardiorespiratory function. The difference between these duration subgroups was statistically significant (P = 0.03), implying that intervention length may have moderated the treatment effect. Nonetheless, heterogeneity within the >12-week subgroup was extremely high (I² = 98%), so the available data did not support the conclusion that longer interventions yielded greater benefits. Interventions with sessions lasting >30 minutes produced a significant effect, whereas those with sessions of ≤30 minutes did not; however, the contrast between the session-duration subgroups was not statistically significant (P = 0.30). Because most trials prescribed exercise at least three times per week at a moderate intensity, there was insufficient variability across studies to identify an optimal training frequency or intensity.

Overall, sports app–based interventions produced beneficial effects on cardiorespiratory function. Intervention duration may represent a key parameter of the exercise prescription that requires further investigation; however, the available evidence was not sufficient to determine an optimal intervention duration, exercise intensity, or session length.

### Effects of Sports Apps on Muscular Fitness Among Adolescents and University Students

This review considered pull-ups in male students and sit-ups in female students as indices of muscular fitness. The meta-analysis indicated that sports app interventions significantly enhanced pull-up performance in male students (MD = 0.74, 95% CI [0.19, 1.28], P = 0.008), although they were accompanied by substantial heterogeneity across studies (I² = 71%). Overall, the results reported by Ting Li [46], Wu and Jiang [44], and Zhang et al. [47]generally favored sports app interventions, whereas Song et al. [43]and some comparisons reported by Zhang et al. [45]showed no clear advantage.

Differences across studies were also reflected in the specific outcomes reported. Ting Li [46] did not observe a statistically significant difference in male pull-up performance between the intervention and control groups. The authors argued that short-term gains in pull-up performance depended not only on overall physical activity levels but also on targeted strength training and movement-technique instruction. Taken together, these results suggested that simply increasing total exercise volume might not have yielded consistent improvements in upper-body muscular performance, and that the app-based delivery of specific training content may have played a critical role.

For female sit-ups, the pooled effect was significant and consistent across studies (MD = 1.99, 95% CI [1.13, 2.86], P < 0.00001; I² = 0%). Ting Li [46] and Wu and Jiang [44] both reported pronounced beneficial effects, and several comparisons presented by Zhang et al. [45] likewise favored the intervention groups. In particular, Ting Li [46] observed a marked improvement in female sit-up performance following the sports app intervention.

Overall, sports app interventions improved students’ muscular fitness; however, the variability observed in male pull-up performance was substantially greater than that seen in female sit-up performance. This discrepancy likely reflected the differing physiological and technical demands of the two tests with respect to body mass, relative strength, muscular endurance, and movement technique. Taken together, these findings supported gains in muscular fitness, but they should not be regarded as direct evidence of increases in maximal muscular strength.

### Effects of Sports Apps on Endurance Running Performance Among Adolescents and University Students

Endurance running performance and sustained exercise capacity were evaluated using the male 1000-m run and the female 800-m run. The meta-analysis indicated that sports app interventions significantly reduced male 1000-m run time (MD = −13.44 s, 95% CI [−25.14, −1.74], P = 0.02), although substantial between-study heterogeneity was present (I² = 87%). Consistent with this overall pattern, Song et al. [43], Wu and Jiang [44], and Zhang et al. [47] reported results that generally favored the intervention groups, and some of these studies documented relatively large performance gains.

Performance outcomes in the female 800-m run were more consistent across studies. Sports app interventions significantly reduced completion time (MD = −9.02 s, 95% CI [−13.40, −4.64], P < 0.0001), and heterogeneity remained relatively low (I² = 32%). Both Song et al. [43]and Wu and Jiang [44] reported pronounced advantages for the sports app intervention. In addition, Wu and Jiang [44] observed enhancements not only in endurance running but also in several other physical fitness indicators among male and female students. On this basis, the authors concluded that a goal-oriented sports app intervention improved University Students’ physical fitness.

Ting Li [46] similarly found significant improvements in the male 1000-m and female 800-m run following the sports app intervention. Participants in the intervention group also engaged more frequently in aerobic and endurance exercise during the intervention. These findings provided a plausible explanation for the pooled results: by recording distance, exercise duration, training tasks, and task completion, sports apps may have supported sustained participation in endurance exercise and thereby improved endurance running performance.

However, performance in the 1000-m and 800-m runs depends not only on local muscular endurance but also on cardiorespiratory function, running economy, and pacing strategy. Therefore, the findings indicated that sports app interventions significantly improved endurance running performance and sustained exercise capacity, rather than providing evidence of improvements in local muscular endurance alone.

### Effects of Sports Apps on Flexibility Among Adolescents and University Students

The meta-analysis of sit-and-reach performance showed that sports app interventions significantly improved flexibility (MD = 2.01 cm, 95% CI [0.91, 3.10], P = 0.0003), although considerable between-study heterogeneity was observed (I² = 81%). Mateo-Orcajada et al. [56], Wu and Jiang [44], and Zhang et al. [45,47]generally reported beneficial effects, whereas Liang and Wang [50], Muntaner-Mas et al. [40], and several comparisons reported by Song et al. [43] yielded more modest gains. Collectively, these results showed that the magnitude of flexibility enhancement varied considerably across studies.

Wu and Jiang[44] reported that participants in the sports app intervention group achieved superior sit-and-reach performance compared with those in the control group, thereby directly supporting the positive pooled effect. In a related study, Liang and Wang [48] found that a 10-week app-supported college physical education intervention improved speed, explosive strength, and flexibility, whereas the effect on endurance was not statistically significant. Taken together, these results indicated that distinct dimensions of physical fitness did not respond uniformly to sports app interventions.

Subgroup analyses indicated significant improvements for interventions lasting both ≤12 weeks (MD = 3.19, 95% CI [1.33, 5.06]) and >12 weeks (MD = 0.71, 95% CI [0.04, 1.38]). The difference between these subgroups was statistically significant (P = 0.01). Notably, the effect estimate was larger for interventions lasting ≤12 weeks than for those extending beyond 12 weeks; thus, the available evidence did not support a linear increase in flexibility gains with longer intervention duration. This pattern likely reflected variation in baseline flexibility, app-based stretching content, and training methods.

#### Overall Interpretation and Research Gaps

Overall, the available evidence has supported beneficial effects of sports app interventions on students’ BMI, cardiorespiratory function, muscular fitness, endurance running performance, and flexibility. The magnitude and consistency of these effects, however, have differed across outcomes. For several indicators, substantial heterogeneity has persisted, implying that participant characteristics, app design features, intervention duration, and exercise dose may have shaped the observed intervention effects. Subgroup analyses have further suggested that intervention duration may have moderated the effects on cardiorespiratory function and flexibility. Nevertheless, the current body of evidence has remained insufficient to determine a universally optimal intervention duration, exercise frequency, exercise intensity, or session duration.

Future studies needed to further elucidate outcome-specific dose–response relationships, standardize the reporting of exercise prescription parameters, and adopt rigorous randomized controlled designs to assess the long-term effects and applicable conditions of different sports app intervention models.

## Limitations

Overall, this systematic review and meta-analysis provided evidence supporting the beneficial effects of sports app interventions on students’ physical fitness. However, several limitations should be acknowledged.

First, the included studies differed in their physical fitness assessment methods and outcome measures. In particular, the measures of cardiorespiratory function, muscular fitness, and endurance running performance were not fully consistent, which may have increased between-study heterogeneity. Second, intervention duration, exercise frequency, exercise intensity, and session duration varied substantially across studies, and some studies incompletely reported these exercise prescription characteristics. Consequently, the available evidence was insufficient to determine an optimal sports app intervention protocol. Finally, this review primarily examined BMI, cardiorespiratory function, muscular fitness, endurance running performance, and flexibility. Skill-related fitness components, such as speed, coordination, and agility, were not systematically evaluated. Therefore, the findings did not fully capture the effects of sports apps on students’ overall physical fitness. Future studies should examine a broader range of outcomes and incorporate variables such as mental health, exercise motivation, and exercise behavior to evaluate the broader health benefits of sports app interventions.

## Conclusions

This systematic review and meta-analysis had several limitations. First, the primary studies employed heterogeneous physical fitness assessment methods and outcome indicators. In particular, the operational definitions and measurement protocols for cardiorespiratory function, muscular fitness, and endurance running performance were not fully aligned across studies, which likely increased between-study heterogeneity.

Second, only a limited number of studies contributed data to several outcomes, and multiple trials exhibited methodological weaknesses in randomization procedures, allocation concealment, and blinding. These deficiencies in study design and execution may have compromised internal validity and, in turn, reduced the robustness of the pooled effect estimates.

Finally, this review primarily examined BMI, cardiorespiratory function, muscular fitness, endurance running performance, and flexibility. Skill-related fitness components, such as speed, coordination, and agility, were not systematically evaluated. Therefore, the findings did not fully capture the effects of sports apps on students’ overall physical fitness. Future studies should examine a broader range of outcomes and incorporate variables such as mental health, exercise motivation, and exercise behavior to evaluate the broader health benefits of sports app interventions.

## Competing Interests

The authors declare that they have no competing interests.

## Funding

The authors received no specific funding for this work.

## Data Availability

The data supporting the findings of this study are available from the corresponding author upon reasonable request.

## Acknowledgments

The authors are grateful to all the authors’ contribution during the completion of this study.

## Supporting information

**S1 Checklist.**

**S1 Table. Physical Fitness-Related Data from the Included Studies.**

**S1 Fig. Subgroup Analysis of BMI by Intervention Duration**

**S2 Fig. Subgroup Analysis of BMI by Exercise Frequency**

**S3 Fig. Subgroup Analysis of BMI by Exercise Intensity**

**S4 Fig. Subgroup Analysis of BMI by Session Duration**

**S5 Fig. Subgroup Analysis of Cardiorespiratory Function by Intervention Duration**

**S6 Fig. Subgroup Analysis of Cardiorespiratory Function by Session Duration**

**S7 Fig. Subgroup Analysis of Sit-and-Reach Performance by Intervention Duration**

## Notes

### Competing Interest Statement

The authors have declared no competing interest.

### Author Declarations

This study is a systematic review and meta-analysis based exclusively on data from previously published studies. No individual participant data were collected, and therefore institutional review board approval and informed consent were not required.

